# Widespread cortical morphological alterations are associated with repetitive blast exposure independent of concussion history

**DOI:** 10.1101/2025.11.19.25340166

**Authors:** Kevin Grant Solar, Nikou Kelardashti, Matthew Ventresca, Rouzbeh Zamyadi, Phillip Johnston, Venkat Bhat, Tom Schweizer, Rakesh Jetly, Anne Wheeler, Jared Rowland, Laura Lester, Sarah Creber, Jing Zhang, Oshin Vartanian, Shawn G Rhind, Benjamin T Dunkley

## Abstract

**Background:** Concussion is a public health crisis, but mounting evidence indicates that repetitive subconcussive head impacts can also lead to lasting structural and functional brain changes. Subconcussive exposures are common in military environments where personnel are routinely subjected to occupational blast overpressure. While functional consequences of blast exposure are increasingly recognized, corresponding structural alterations remain less consistently reported. Here, we investigated how cumulative subconcussive exposure relates to cortical and subcortical brain structure in a military cohort.

**Methods:** Using FreeSurfer, we analyzed high-resolution T1-weighted images from 80 participants (n = 41 high blast, n = 39 low blast; 4 and 8 females, respectively), grouped by lifetime blast exposure history using the generalized blast exposure value. Vertex-wise general linear models tested for cortical differences in cortical volume, thickness, surface area, and curvature, adjusting for age, trauma exposure, and diagnosed concussion count. ANCOVAs evaluated subcortical volume differences.

**Results:** Compared to the low blast group, individuals with high blast exposure demonstrated widespread cortical alterations. Across metrics, higher values were predominantly observed in frontal and central regions, including the precentral and superior frontal gyri, whereas lower metrics mainly appeared in temporal, parietal, and occipital cortices. Specifically, high blast participants showed greater cortical volume, thickness, and surface area in precentral and superior frontal regions, higher curvature in frontoparietal and temporal areas, and reduced volume, surface area, and thickness in lateral temporal, parietal, and occipital cortices (e.g., supramarginal, inferior parietal, and lingual gyri) and the entorhinal cortex. Subcortically, high blast participants showed no volumetric differences from the low blast group.

**Conclusion:** These findings provide evidence that repetitive subconcussive blast exposure alters cortical morphometry, independent of concussion history, suggesting complex, spatially heterogenous structural changes and highlighting the need for proactive monitoring and mitigation strategies in operational populations.

## 1. Background

Blast overpressure is a pervasive occupational hazard in military settings, arising from repeated exposure to pressure waves generated by explosives and heavy weapons, even when no direct head impact occurs [1]. While subconcussive blast exposures are often overshadowed by clinically diagnosed concussion or traumatic brain injury (TBI), evidence indicates that cumulative blast exposure can produce lasting neurobiological consequences independent of concussion history [2–6]. Parallel insights from chronic traumatic encephalopathy (CTE), a progressive tauopathy diagnosed postmortem, suggest that cumulative head impact burden, rather than diagnosed concussion history, is a superior predictor of disease severity [7, 8], although the underlying pathogenic mechanisms remain poorly understood [9, 10]. Neuropathological hallmarks of cumulative blast exposure or repetitive mild head impacts include astrogliosis at the cortex–white matter junction [11, 12]. However, among military veterans, CTE pathology is less consistently detected than in civilians with non-military TBI, suggesting differences in underlying mechanisms [13, 14].

Since CTE and other proteinopathic dementias can only be definitively diagnosed postmortem, identifying reliable *in vivo* predictive biomarkers is essential for advancing early diagnosis and therapeutic intervention [15]. Neuroimaging studies have begun to reveal early *in vivo* cortical and subcortical changes associated with repeated subconcussive impacts, showing mixed effects [16–19]. For instance, a longitudinal analysis found that collegiate football players experienced less cortical thinning with age relative to non-contact athletes (volleyball players) [20]. A longitudinal study of high school football athletes, following a single season of play, reported that greater exposure to high-magnitude head impact, measured via helmet sensors, predicted cortical thinning in sensorimotor and frontal cortices, regardless of concussion status [21]. Similarly, a recent cross-sectional study of former collegiate and professional American football players reported reduced cortical thickness and volumes, including in the amygdala, entorhinal cortex, and superior frontal gyrus, relative to unexposed controls [22].

In military populations, cross-sectional studies have shown that Veterans with both concussion and blast exposure exhibit greater cortical thinning in regions such as the frontal gyrus and orbitofrontal cortex compared to those with concussion alone [23]. Vartanian et al. found that in military breachers repeatedly exposed to low-level blast, voxel-based morphometry revealed reduced gray matter volume in the right superior frontal gyrus compared to controls, suggesting cumulative structural brain alterations [24]. Similarly, cortical thinning of the anterior cingulate and superior frontal cortex has been reported in service members with blast-related concussion relative to those with non-blast concussion [25]. More recently, findings from a large longitudinal military cohort (n=774), demonstrated that a history of blast-related concussion was associated with reduced white matter and subcortical gray matter volumes, which in turn correlated with slower processing speed and poorer working memory [26]. Despite these advances, prior morphometric studies of blast-related subconcussion have primarily focused on cortical volume and thickness [26–28]. A comprehensive, high-resolution, surface-based morphometry assessment incorporating cortical volume, thickness, surface area, curvature, alongside subcortical structures, may offer greater sensitivity to subtle and spatially distributed structural changes, distinguishing subconcussive effects from those of diagnosed concussions or mild traumatic brain injury (mTBI).

In this study, we applied the FreeSurfer pipeline to examine cortical and subcortical morphometry in Canadian Armed Forces (CAF), Royal Canadian Mounted Police (RCMP), and Veteran personnel. Participants were stratified into high and low blast exposure groups using a median split on the Generalized Blast Exposure Value (GBEV) [29] as described in prior work [30, 31], with analyses that controlled for concussion history, age, and psychological trauma. We hypothesized that individuals with higher cumulative blast exposure would exhibit reduced cortical volume and thinning in frontal, parietal, and temporal regions, along with alterations in surface area and curvature, as well as region-specific changes in subcortical volume. By delineating *in vivo* structural signatures of repetitive subconcussive blast exposure, this research aims to advance early detection, prevention, and intervention strategies that safeguard the health and performance of vulnerable operational populations.

## 2. Materials and methods

### 2.1 Participants

Participants were enrolled as part of a large, multidimensional study designed to investigate brain and mental health in military populations using both cross-sectional and longitudinal approaches. Recruitment efforts targeted current and former members of the Canadian Armed Forces (CAF) and Royal Canadian Mounted Police (RCMP), using outreach through military units, health services, and military-related organizations (e.g., Project Enlist, Veterans Affairs Canada, and other Veteran support networks).

Inclusion criteria required participants to be current or former CAF/RCMP personnel with a history of occupational exposure to blast, possess sufficient English proficiency and cognitive ability to complete the study procedures, and have no contraindications to MRI or MEG scanning. Data collection occurred between September 2020 and May 2023 at the Hospital for Sick Children in Toronto, Ontario, Canada.

In total, 84 participants were recruited; 4 participants did not complete MRI. For all MRI analyses, 80 CAF/RCMP personnel and Veterans from various backgrounds, including personnel from combat and emergency response roles such as weapons operation, armoured units, engineering, explosive disposal, and artillery, were divided into two groups based on a median split of blast exposure levels assessed by the GBEV questionnaire: *n* = 41 high blast exposure group (aged 26.4 to 65.7 years; mean 48.1 ± 9.0 years; 4 females) and *n* = 39 low blast exposure group (aged 28.0 to 63.3 years; mean 48.1 ± 9.4 years; 8 females) (see Figure 1B in Solar et al. 2023 for group distributions) [31]. The GBEV is a tool for standardizing a lifetime of blast exposure by quantifying units of blast exposure across five categories of blast type/severity (i.e., Small Arms, Large Arms, Artillery, Small Explosives, Large Explosives) [29]. The resulting GBEV score serves as a comprehensive measure of blast exposure history, facilitating the assessment of potential health risks associated with repeated low-level blast events. This study was approved by the Hospital for Sick Children Research Ethics Board and Defence Research and Development Canada’s (DRDC) Human Research Ethics Committee. Written informed consent was obtained from all participants in accordance with the Declaration of Helsinki.

**Figure 1.**
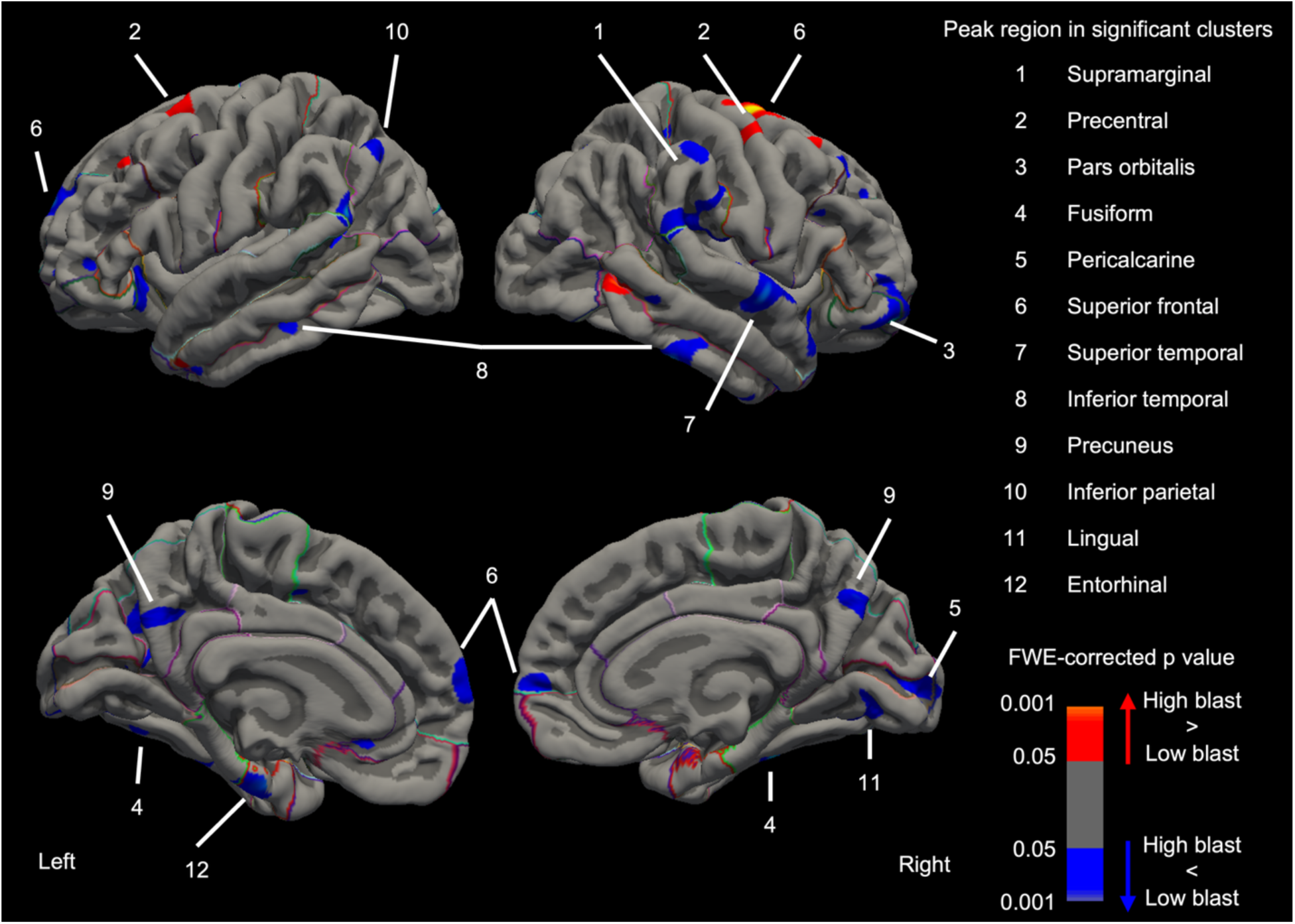
Bilateral cortical volume differences with higher blast exposure. Compared to low blast exposure, high blast exposure was associated with regional reductions (blue) in cortical volume, alongside focal volume increases in frontal nodes. Colours indicate the direction and significance of group differences.

### 2.2 Neurobehavioural and psychiatric outcomes

All participants completed neurobehavioural and mental health assessments which included: Generalized Anxiety Disorder 7 screener (GAD7); Patient Health Questionnaire (PHQ9); PTSD Checklist Military Version (PCL-M); and the neurobehavioural symptom evaluation from the Sports Concussion Assessment Tool 5 (SCAT5) [32]. Participants also completed brief evaluations related to diagnosed lifetime concussion history, using the Acute Concussion Evaluation (ACE) [33], and defined by the US Department of Defence/Veterans Affairs guidelines for mTBI as injury resulting in loss of consciousness <30 minutes, post-traumatic amnesia <24 hours, and a Glasgow Coma Score of 13 or more. Psychological trauma as traumatic stress history was also estimated with the Brief Trauma Questionnaire (BTQ) [34].

### 2.3 MRI acquisition and processing

High-resolution T1-weighted anatomical MRI scans were acquired on a 3T Siemens PrismaFit scanner using a 20-channel head and neck coil at the Hospital for Sick Children in Toronto, Ontario, Canada. A 3D magnetization-prepared rapid gradient echo (MPRAGE) sequence was utilized with 0.8 mm isotropic resolution in 5:01 minutes (repetition time (TR) = 1870 ms, echo time (TE) = 3.1 ms, inversion time (TI) = 945 ms, field of view (FOV) = 240 × 256 mm, with 240 sagittal slices).

T1-weighted images were processed using FreeSurfer version 7.4 [35], which performs automated reconstruction of cortical surfaces and segmentation of subcortical structures via the recon-all pipeline. This includes motion correction, brain extraction, intensity normalization, Talairach registration, surface tessellation, topology correction, and cortical parcellation using the Desikan-Killiany atlas.

### 2.4 Statistical analysis

Given the known influence of age, sex, concussion history, and psychological trauma on brain structure, these variables were considered as covariates.

#### 2.4.1 Participant demographics and outcomes

Analyses of covariance (ANCOVA) in JASP version 0.18.0 (JASP, 2023) were used to test for blast-related group differences in outcome measures (GAD7, PHQ9, PCL-M, SCAT5) while adjusting for age, sex, traumatic stress history (BTQ), and number of diagnosed concussions (ACE). ANCOVA was also used to test for group differences in outcomes, stratified by concussion/no concussion history, while adjusting for age and sex – these results are available in a prior study published on the same cohort [31].

#### 2.4.2 Cortical morphometry analysis

Statistical analyses of cortical morphology were conducted in FreeSurfer version 7.4 [35], using multiple linear regression to estimate the effects of blast exposure group on grey matter measures. FreeSurfer was used to extract vertex-wise measures of cortical thickness, surface area, volume, and mean curvature. Cortical thickness was calculated as the distance between the white matter and pial surfaces, while surface area and curvature were computed from the tessellated white matter surface. Cortical volume at each vertex was derived by multiplying thickness by surface area. These morphometric measures were resampled to the fsaverage spherical atlas, which enables cross-subject comparisons by aligning cortical folding patterns (sulci and gyri) to a common surface template.

Group-level statistical analyses were performed using FreeSurfer’s general linear modeling tool (mri_glmfit) to examine the effects of cumulative blast exposure on each cortical metric. Covariates included age, history of psychological trauma (BTQ), and number of diagnosed concussions (ACE). Sex was not included as a covariate due to insufficient representation in the high blast exposure group (n=4 females), which did not meet FreeSurfer’s requirements for including multiple covariates in models with sparse group membership. Cluster-wise correction for multiple comparisons was performed using permutation-based family-wise error (FWE) control with PALM (Permutation Analysis of Linear Models). Vertex-wise clusters were initially formed using a threshold of –log₁₀(p) = 1.3 (corresponding to p = 0.05, uncorrected). Non-parametric statistical significance was determined using 1000 permutations, with clusters considered significant if they survived the FWE-corrected p < 0.05 threshold. The minimum cluster size was empirically determined from the permutation-based null distribution.

#### 2.4.3 Subcortical volume region-of-interest (ROI) analysis

Subcortical structures were segmented using FreeSurfer’s automated labeling system (aseg.mgz), which assigns regions based on voxel intensity and probabilistic anatomical priors [35]. ROIs included the thalamus, caudate, putamen, pallidum, hippocampus, amygdala, and nucleus accumbens, subcortical regions implicated in cognitive, emotional, and motor processes that are vulnerable to diffuse injury from blast exposure. High versus low blast exposure group comparisons were conducted using ANCOVAs in JASP with age, trauma history (BTQ), number of diagnosed concussions (ACE), and estimated total intracranial volume (eTIV; to account for individual differences in head size) as covariates. As in the cortical analysis, sex was excluded due to insufficient representation. Multiple comparisons were corrected using the false discovery rate (FDR) method [36].

## 3. Results

### 3.1 Demographics

The age of the high blast group did not differ significantly from the low blast group (high blast group *n* = 41, 48.1 ± 9.0 [26.4-65.7] years; low blast group *n* = 39, 47.8 ± 9.5 [28.0-63.3] years; F(1,78) = 1.9E-4, p = 0.9), nor did sex ratios (high blast group 37 males, 4 females; low blast group 32 males, 8 females; F(1,78) = 1.8, p = 0.2), number of concussions (high blast group mean = 2.0; low blast group mean = 1.3; F(1,78) = 1.9, p = 0.2), or psychological trauma indexed by the BTQ (high blast group mean = 5.7; low blast group mean = 5.4; F(1,78) = 0.1, p = 0.7).

### 3.2 Widespread cortical alterations in high blast exposure

Significant group differences in cortical morphometry were observed between individuals with high versus low cumulative blast exposure, after correction for multiple comparisons (cluster-wise FWE-corrected p < 0.05). Regionally divergent morphometric patterns were observed in the high blast group. Higher values observed across cortical volume, thickness, surface area, and curvature were largely concentrated in frontal and central regions, particularly the precentral and superior frontal gyri, while lower values were most prominent in lateral temporal, parietal, and occipital cortices, including the supramarginal, inferior parietal, lingual, and entorhinal regions. These findings suggest complex, spatially heterogeneous cortical changes associated with repetitive subconcussive blast exposure.

#### 3.2.1 Cortical volume

Compared to the low blast group, participants in the high blast group showed significantly greater cortical volume in clusters located in the precentral gyrus and superior frontal gyrus. In contrast, the high blast group exhibited lower cortical volume in the supramarginal gyrus, pars orbitalis, fusiform gyrus, pericalcarine cortex, superior temporal gyrus, inferior temporal gyrus, precuneus, inferior parietal lobule, lingual gyrus, and entorhinal cortex **(Figure 1)**.

#### 3.2.2 Cortical thickness

The high blast group demonstrated significantly greater cortical thickness in the supramarginal gyrus, precentral gyrus, postcentral gyrus, fusiform gyrus, pericalcarine cortex, middle temporal gyrus, lateral occipital cortex, precuneus, superior parietal lobule, inferior parietal lobule, and anterior cingulate cortex. Reduced thickness in the high blast group was observed in the superior frontal gyrus **(Figure 2).**

**Figure 2.**
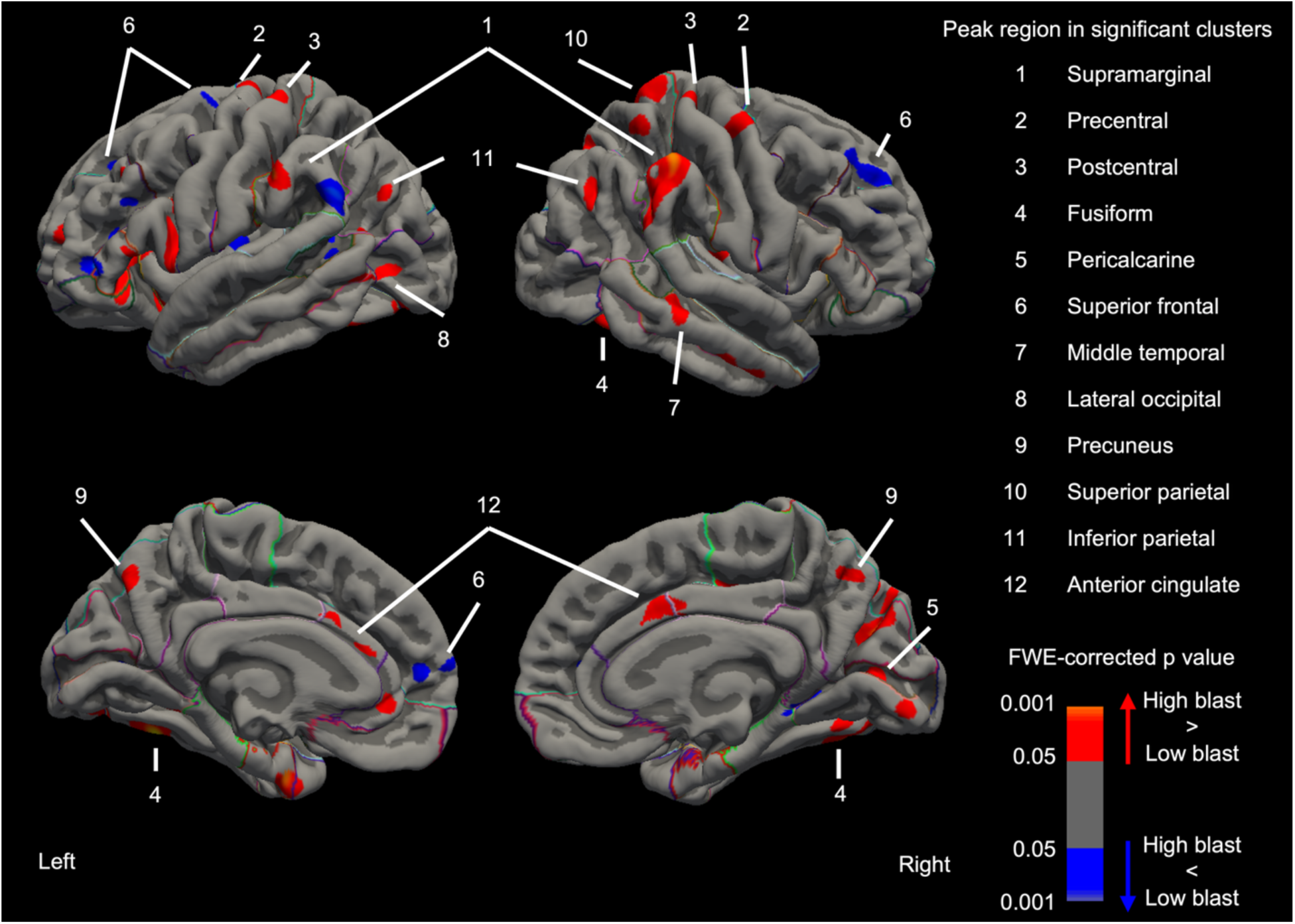
High blast exposure presents with increased cortical thickness (red), with a smaller number of areas presenting with reduced cortical thickness (blue). Colours indicate the direction and significance of group differences.

#### 3.2.3 Cortical surface area

Significant increases in cortical surface area were identified in the high blast group within the precentral gyrus, middle frontal gyrus, lateral orbitofrontal cortex, and superior frontal gyrus. Conversely, reduced surface area in the high blast group was seen in the lingual gyrus, postcentral gyrus, superior temporal gyrus, middle temporal gyrus, precuneus, superior parietal lobule, inferior parietal lobule, and entorhinal cortex **(Figure 3).**

**Figure 3.**
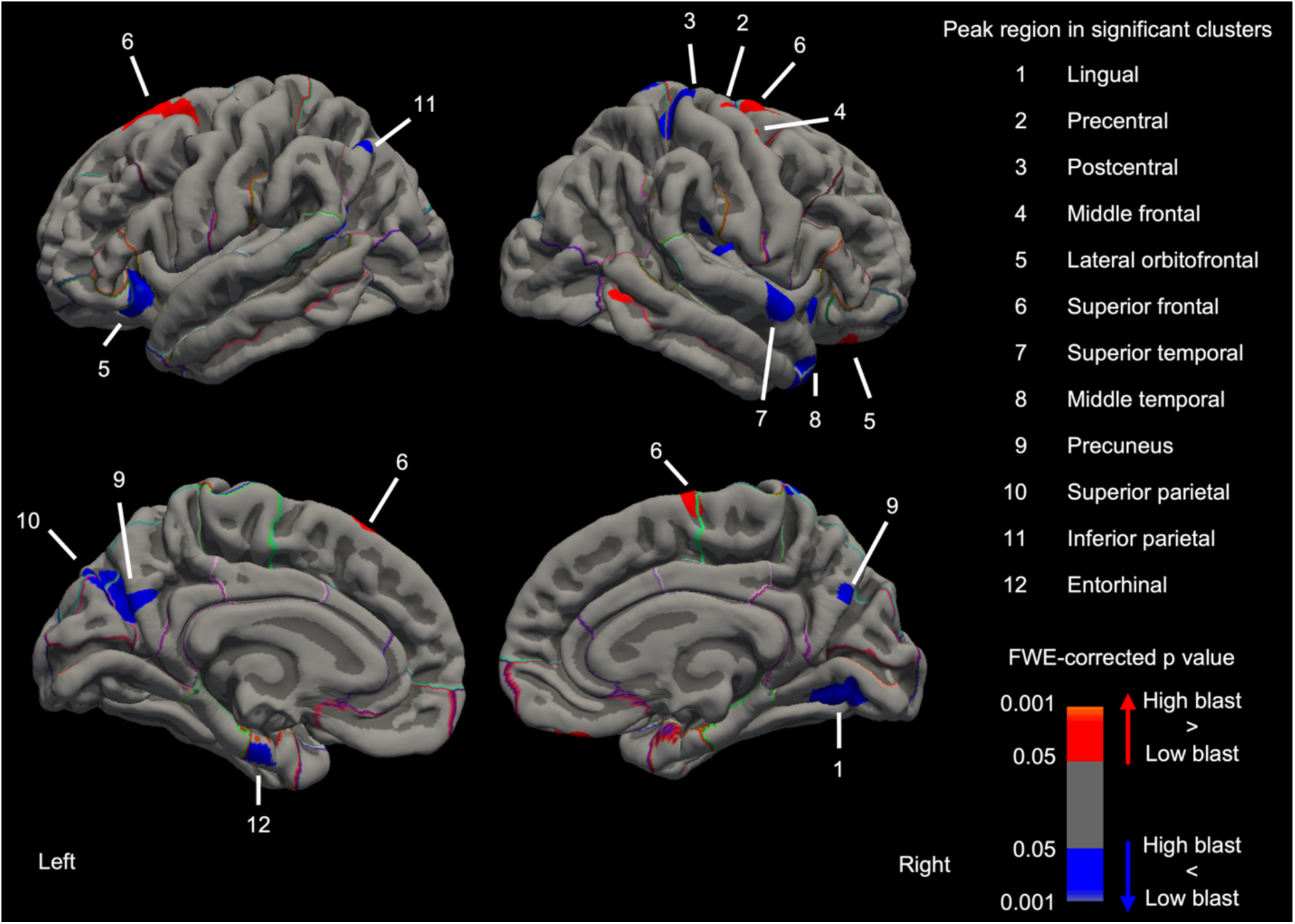
High blast exposure was associated with mainly reduced cortical surface area (blue) with focal higher surface area in superior frontal and precentral gyri (red). Colours indicate the direction and significance of group differences.

#### 3.2.4 Cortical curvature

Participants with high blast exposure exhibited significantly greater mean curvature in the paracentral lobule, supramarginal gyrus, superior frontal gyrus, superior temporal gyrus, middle temporal gyrus, precuneus, medial orbitofrontal cortex, and lateral occipital cortex. In contrast, lower cortical curvature in the high blast group was observed in the precentral gyrus, postcentral gyrus, middle frontal gyrus, and inferior parietal lobule **(Figure 4)**.

**Figure 4.**
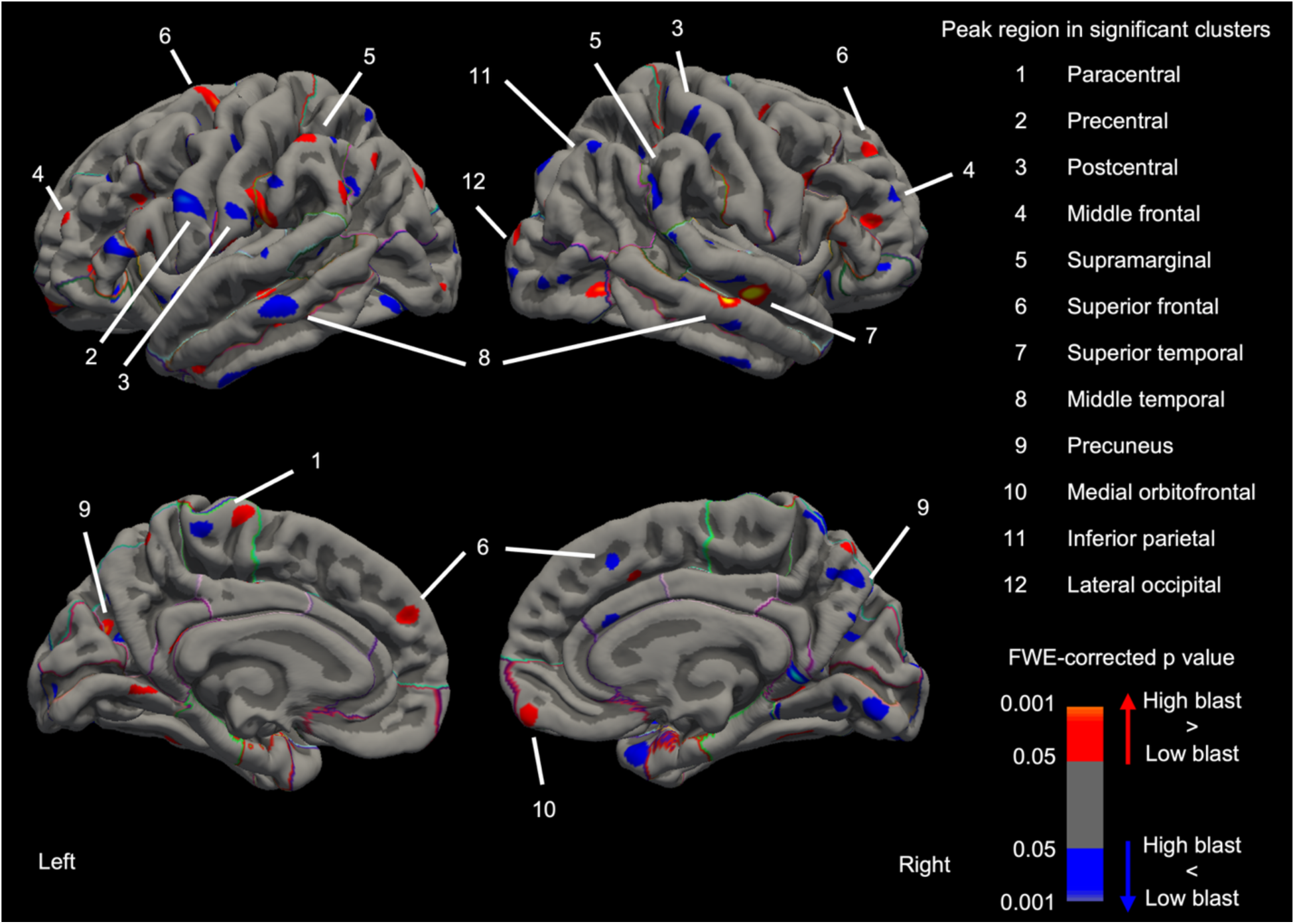
Individuals with high blast exposure showed both increases (red) and decreases (blue) in cortical curvature across widespread regions. Colours indicate the direction and significance of group differences.

### 3.3 No subcortical alterations in high blast exposure

Analysis of subcortical volumes revealed no significant group differences related to cumulative blast exposure after the FDR correction. Uncorrected trends did not survive correction, indicating that subcortical effects were weaker and less spatially consistent than the widespread cortical alterations.

## 4. Discussion

### 4.1 Summary

This study provides novel evidence that repetitive subconcussive blast exposure is associated with widespread, regionally heterogeneous cortical alterations in military personnel and Veterans, independent of concussion or trauma history. Using surface-based morphometry and ROI-based volumetric analyses, high cumulative blast exposure was characterized by higher cortical volume, thickness, and surface area in frontal and central regions, particularly the precentral and superior frontal gyri, alongside higher cortical curvature in frontoparietal and temporal areas. In contrast, lower cortical volume, surface area, and thickness were observed in lateral temporal, parietal, and occipital regions, including the supramarginal, inferior parietal, and lingual gyri and the entorhinal cortex. Subcortical analyses revealed no significant differences in the high blast group. These findings align with and extend prior work in concussion and blast-related mTBI, demonstrating that even in the absence of clinically diagnosed TBI, cumulative subconcussive exposure can elicit measurable structural brain alterations.

The distributed and multimodal pattern of these alterations supports a diffuse injury model and raises critical questions regarding the long-term neurobiological consequences of repeated low-level blast exposure in occupational settings.

### 4.2 Widespread cortical volume alterations in individuals with high blast exposure

The widespread cortical volume reductions in the high blast group, compared to the low blast individuals, spanned the temporal, parietal, occipital, and limbic cortices.

This aligns with previous studies that have linked repetitive head impacts to structural degeneration, including neuronal loss and dendritic retraction [37]. In most affected areas, the decreases in volume occurred alongside contraction of surface area but preserved or increased cortical thickness. This suggests that the primary driver of reduced cortical volume was the loss of surface area and not cortical thinning.

In contrast, a subset of frontal regions showed increases in volume. The precentral gyrus showed a volume increase in the high blast group, while the superior frontal gyrus showed mixed results. These focal increases in volume may reflect neuroinflammation rather than neuronal hypertrophy [38, 39]. Postmortem studies in individuals with repetitive head impact exposure have reported increased astrogliosis at the grey-white matter interface in the dorsolateral frontal cortex [11], suggesting that frontal volume increase seen in our study may reflect underlying reactive gliosis [40].

### 4.3 Increased cortical thickness in high blast exposure group

In contrast to volume findings, cortical thickness was predominantly increased in the high blast group compared to low blast individuals. Although cortical thickness is mostly reported to decrease post-blast exposure, the increased thickness that we see may reflect early neurobiological responses such as inflammation, reactive gliosis, or reduced cortical pruning [41]. Increased cortical thickness patterns have been reported in contact-sport athletes with a history of concussion [20].

Notably, the anterior cingulate cortex also showed increased thickness, most likely reflecting its anatomical vulnerability due to its position near blast wave entry point via the orbits and presence of von Economo neurons which are metabolically demanding and sensitive to mechanical stress. This finding aligns with prior work showing increased thickness in ACC being associated with higher blast exposure [12].

These findings are consistent with those of Diociasi *et al*., who reported that repetitive blast exposure in Special Operations Forces personnel was associated with distinct alterations in resting-state functional connectivity and cortical morphology – most notably increased cortical volume in the lateral occipital cortex among high-exposure individuals compared with both healthy controls and low-exposure peers [42]. These volumetric and connectivity changes within occipital, frontal, and parietal networks correlated with greater neurobehavioral symptom severity, suggesting that alterations in functional connectivity may represent early and sensitive indicators of blast-related brain injury, even in the absence of overt structural abnormalities.

By contrast, the superior frontal gyrus represents an exception, showing reduced thickness despite increased surface area and curvature (see Subsection 4.4). This is in line with other studies showing greater frontal cortical thinning [23, 43].

### 4.4 Altered surface area and cortical curvature in high blast exposure

In the high blast group, surface area was predominantly reduced across occipital, temporal, and parietal regions – a notable finding considering that surface area is rarely reported in blast injury studies [26–28]. In many of these regions, volume loss co-occurred with either preserved or increased cortical thickness, suggesting that surface area contraction, rather than thinning, was the principal driver of volumetric decline. This pattern aligns with aging studies showing that cortical thickness and surface area follow partly independent trajectories and are shaped by distinct genetic and biological influences [44, 45], and can diverge dynamically across adulthood [46]. Whereas aging is often characterized by a “cortical stretching” pattern, thickness decreases with relative preservation or expansion of surface area [44], here, we show that blast exposure largely results in reduced surface area with preserved or increased thickness, suggesting a morphometric response distinct from typical aging processes.

The superior frontal gyrus again represented an exception, showing higher surface area accompanied by reduced thickness, and higher curvature. This pattern resembles the cortical stretching profile observed in aging [44]. Elevated curvature may reflect cortical restructuring or altered folding dynamics secondary to white-matter disruption or loss of mechanical tension, consistent with findings of widespread curvature increases in veterans with mTBI [47, 48]. These results underscore the idea that cortical metrics – surface area, thickness, and curvature – reflect distinct yet interacting neurobiological processes and may diverge regionally depending on local mechanical and connective vulnerabilities to blast-related injury.

### 4.5 No subcortical alterations in high blast exposure

Although previous studies of blast exposure and mild TBI have reported subcortical alterations [16–19, 26], particularly within limbic and basal ganglia structures, our results did not reveal significant group differences after correcting for multiple comparisons. This suggests that cumulative subconcussive blast exposure may primarily affect cortical morphometry rather than gross subcortical volumes, at least within the sensitivity limits of the current sample. It is possible that subcortical alterations emerge only with higher exposure intensity, longer post-exposure intervals, or through microstructural changes not captured by volumetric measures but by diffusion-weighted MRI which has revealed mixed results, including changes in the hippocampus and thalamus [49].

### 4.6 Future directions

A key strength of this paper is the analysis of surface-based morphometry across multiple metrics. This allowed for a nuanced understanding of cortical remodeling in high blast exposure. Through adjusting for concussion history, these results represent the unique contribution of blast exposure. However, there are several limitations associated with this study. First, blast exposure was measured using the subjective self-reported probe of lifetime blast exposure with the GBEV scale, which may introduce recall bias; objective blast exposure data, such as lifetime pressure gauge use, would be advantageous, but due to practical limitations are not feasible to capture in our current cohort. Second, high-level (incoming) and low-level (outgoing) blasts differ. High-level events are typically single war-zone incidents, whereas low-level exposures accumulate during training [50]. Although we focus on repeated low-level (subconcussive) blast exposure, unmeasured prior high-level exposures, particularly from past deployments, may confound our findings. Third, the sex imbalance in our sample limits the generalizability of the findings, highlighting the need for future research to focus on structural and functional brain changes following blast exposure in female participants. Fourth, the cross-sectional design of this study hinders understanding of the temporal trajectory of the brain changes. Finally, while morphometric measures are sensitive to structural differences, we cannot fully understand the underlying mechanism – neuronal loss, neuroinflammation, white matter injury – for the observed effects with structural data in isolation. Therefore, future studies should collect longitudinal data and combine structural, functional, and molecular imaging to establish causal mechanisms and track the progression of neural changes.

## 5. Conclusion

In conclusion, high blast exposure, independent of concussion history, is associated with widespread alterations in cortical morphometry. Overall, individuals with high cumulative blast exposure exhibited reduced cortical volume and surface area alongside increased cortical thickness, suggesting that surface area loss, rather than cortical thinning, is the predominant contributor to volume reduction. Notably, several frontal regions demonstrated morphometric patterns consistent with inflammatory remodeling, distinct from those typically observed in normal aging or age-related neurodegenerative disease, potentially reflecting blast-specific neurotraumatic processes.

These findings highlight the importance of evaluating multiple morphometric indices to comprehensively characterize blast-related brain alterations and their potential utility as biomarkers for early detection, longitudinal monitoring, and targeted intervention in at-risk military populations. Moreover, the results emphasize the critical need to extend research beyond clinically diagnosed concussions to include individuals exposed to repetitive “subconcussive” blasts, who may face cumulative and underrecognized neurobiological effects.

## List of abbreviations

ACE: Acute Concussion Evaluation
ACC: Anterior Cingulate Cortex
ANCOVA: Analysis of Covariance
BTQ: Brief Trauma Questionnaire
CAF: Canadian Armed Forces
CTE: Chronic Traumatic Encephalopathy
eTIV: Estimated Total Intracranial Volume
FDR: False Discovery Rate
FOV: Field of View
FWE: Family-Wise Error
GAD7: Generalized Anxiety Disorder 7 Screener
GBEV: Generalized Blast Exposure Value
GLM: General Linear Model
JASP: Jeffrey’s Amazing Statistics Program
mTBI: Mild Traumatic Brain Injury
MEG: Magnetoencephalography
MPRAGE: Magnetization-Prepared Rapid Gradient
Echo MRI: Magnetic Resonance Imaging
PCL-M: PTSD Checklist: Military Version
PHQ9: Patient Health Questionnaire-9
PTSD: Post-Traumatic Stress Disorder
ROI: Region of Interest
RCMP: Royal Canadian Mounted Police
SCAT5: Sports Concussion Assessment Tool: 5th Edition
T1w: T1-weighted (MRI sequence)
TBI: Traumatic Brain Injury
TE: Echo Time
TI: Inversion Time
TR: Repetition Time

## Availability of data and materials

Defence considerations related to the confidential nature of the human data collected means it is not publicly available.

## Acknowledgements

We have no acknowledgements beyond the funding information below.

## Ethics approval

This study was approved by the Hospital for Sick Children Research Ethics Board and Defence Research and Development Canada’s Human Research Ethics Committee and conducted in accordance with the Declaration of Helsinki.

## Consent to participate

Written informed consent was obtained from all participants.

